# Synaptic processes and immune-related pathways implicated in Tourette Syndrome

**DOI:** 10.1101/2020.04.24.20047845

**Authors:** Fotis Tsetsos, Dongmei Yu, Jae Hoon Sul, Alden Y. Huang, Cornelia Illmann, Lisa Osiecki, Sabrina M. Darrow, Matthew E. Hirschtritt, Erica Greenberg, Kirsten R. Muller-Vahl, Manfred Stuhrmann, Yves Dion, Guy A. Rouleau, Harald Aschauer, Mara Stamenkovic, Monika Schlögelhofer, Paul Sandor, Cathy L. Barr, Marco A. Grados, Harvey S. Singer, Markus M. Nöthen, Johannes Hebebrand, Anke Hinney, Robert A. King, Thomas V. Fernandez, Csaba Barta, Zsanett Tarnok, Peter Nagy, Christel Depienne, Yulia Worbe, Andreas Hartmann, Cathy L. Budman, Renata Rizzo, Gholson J. Lyon, William M. McMahon, James R. Batterson, Danielle C. Cath, Irene A. Malaty, Michael S. Okun, Cheston Berlin, Douglas W. Woods, Paul C. Lee, Joseph Jankovic, Mary M. Robertson, Donald L. Gilbert, Lawrence W. Brown, Barbara J. Coffey, Andrea Dietrich, Pieter J. Hoekstra, Samuel Kuperman, Samuel H. Zinner, Michael Wagner, James A Knowles, A. Jeremy Willsey, Jay A. Tischfield, Gary A. Heiman, Nancy J. Cox, Nelson B. Freimer, Benjamin M. Neale, Lea K. Davis, Giovanni Coppola, Carol A. Mathews, Jeremiah M. Scharf, Peristera Paschou, on behalf of the Tourette Association of America International Consortium for Genetics, the Gilles de la Tourette GWAS Replication Initiative, the Tourette International Collaborative Genetics Study, and the Psychiatric Genomics Consortium Tourette Syndrome Working Group

## Abstract

Tourette Syndrome (TS) is a neuropsychiatric disorder of complex genetic architecture involving multiple interacting genes. Here, we sought to elucidate the pathways that underlie the neurobiology of the disorder through genome-wide analysis. We analyzed genome-wide genotypic data of 3581 individuals with Tourette Syndrome (TS) and 7682 ancestry-matched controls and investigated associations of TS with sets of genes that are expressed in particular cell types and operate in specific neuronal and glial functions. We employed a self-contained, set-based association method (SBA) as well as a competitive gene set method (MAGMA) using individual-level genotype data to perform a comprehensive investigation of the biological background of TS. Our SBA analysis identified three significant gene sets after Bonferroni correction, implicating Ligand-gated Ion Channel Signaling, Lymphocytic, and Cell Adhesion and Trans-synaptic Signaling processes. MAGMA analysis further supported the involvement of the Cell Adhesion and Trans-synaptic Signaling gene set. The Lymphocytic gene set was driven by variants in *FLT3*, raising an intriguing hypothesis for the involvement of a neuroinflammatory element in TS pathogenesis. The indications of involvement of Ligand-gated Ion Channel Signaling reinforce the role of GABA in TS, while the association of Cell Adhesion and Trans-synaptic Signaling gene set provides additional support for the role of adhesion molecules in neuropsychiatric disorders.

## Introduction

Tourette Syndrome (TS) is a chronic neurodevelopmental disorder characterized by several motor tics and at least one vocal tic that persist more than a year [1]. Its prevalence is between 0.6-1% in school-aged children [2, 3]. Although TS is highly polygenic in nature, it is also highly heritable [4]. The population-based heritability is estimated at 0.7 [5, 6], with SNP-based heritability ranging from 21% [7] to 58% [4] of the total. The genetic risk for TS that is derived from common variants is spread throughout the genome [4]. The two genome-wide association studies (GWAS) conducted to date [7, 8] suggest that TS genetic variants may be associated, in aggregate, with tissues within the cortico-striatal and cortico-cerebellar circuits, and in particular, the dorsolateral prefrontal cortex. The GWAS results also demonstrated significant ability to predict tic severity using TS polygenic risk scores [7, 9]. A genome-wide CNV study identified rare structural variation contributing to TS on the *NRXN1* and *CNTN6* genes [10]. De novo mutation analysis studies in trios have highlighted two high confidence genes, *CELSR* and *WWC1*, and four probable genes, *OPA1, NIPBL, FN1*, and *FBN2* to be associated with TS [11, 12].

Investigating clusters of genes, rather than relying on single marker tests is an approach that can significantly boost power in a genomewide setting [13]. This approach has already proven useful in early genome-wide studies of TS. The first published TS GWAS, which included 1285 cases and 4964 ancestry-matched controls did not identify any genome-wide significant loci. However, by partitioning functional- and cell type-specific genes into gene sets, an involvement of genes implicated in astrocyte carbohydrate metabolism was observed, with a particular enrichment in astrocyte-neuron metabolic coupling [14]. Here, we investigated further the pathways that underlie the neurobiology of TS, performing gene set analysis on a much larger sample of cases with TS and controls from the second wave TS GWAS. We employed both a competitive gene set analysis as implemented through MAGMA, as well as a self-contained analysis through a set-based association method (SBA).

## Methods and Materials

### Samples and quality control

The sample collection has been extensively described previously [7, 8]. IRB approvals and consent forms were in place for all data collected and analyzed as part of this project. For the purposes of our analysis, we combined 1285 cases with TS and 4964 ancestry-matched controls from the first wave TS GWAS, with 2918 TS cases and 3856 ancestry-matched controls from the second wave TS GWAS. Standard GWAS quality control procedures were employed [15, 16]. The data were partitioned first by genotyping platform and then by ancestry. The sample call rate threshold was set to 0.98, and the inbreeding coefficient threshold to 0.2. A marker call rate threshold was defined at 0.98, case-control differential missingness threshold at 0.02, and Hardy-Weinberg Equilibrium (HWE) threshold to 10^*−*6^ for controls and 10^*−*10^ for cases. Before merging the partitioned datasets, we performed pairwise tests of association and missingness between the case-only and control-only subgroups to address potential batch effect issues. All SNPs with p-values *≤* 10^*−*06^ in any of these pairwise quality control analyses were removed. After merging all datasets, Principal Component Analysis was utilized to remove samples that deviated more than 6 standard deviations and to ensure the homogeneity of our samples in the ancestry space of the first 10 principal components, through the use of the EIGENSOFT suite [17]. Identity-by-descent analysis with a threshold of 0.1875 was used to remove related samples, and thus to avoid confounding by cryptic relatedness. After quality control, the final merged dataset consisted of 3581 cases with TS and 7682 ancestry-matched controls on a total of 236,248 SNPs, annotated using dbSNP version 137 and the hg19 genomic coordinates.

We assessed the genomic variation in our data through PCA analysis to identify potential population structure. The variation in our data was reduced to a triangular shape in the two-dimensional space of the first two principal components. One tip was occupied by Ashkenazi Jewish samples, the second by the Southern European samples and the other by the North Europeans. Depicting geography, the Southern to Nothern axis was populated by European-ancestry samples. The first five principal components were added to the association model as covariates, in order to avoid population structure influencing our results.

### Gene sets

We collected neural-related gene sets from multiple studies on pathway analyses in neuropsychiatric disorders [14, 18–22]. These studies relied on an evolving list of functionally-partitioned gene sets, focusing mainly on neural gene sets, including synaptic, glial sets, and neural cell-associated processes. We added a lymphocytic gene set also described in these studies [21], in order to also investigate potential neuro-immune interactions.

In total, we obtained 51 gene sets, which we transcribed into NCBI entrez IDs and subsequently filtered by removing gene sets that contained fewer than 10 genes. 45 gene sets fit our criteria and were used to conduct the analyses.

We examined two primary categories of pathway analysis methods, the competitive [23] and the self-contained test [24]. The competitive test compares the association signal yielded by the tested gene set to the association signals that do not reside in it [23, 25]. In this type of test, the null hypothesis is that the tested gene set attains the same level of association with disease as equivalent random gene sets. In contrast, the self-contained test investigates associations of each tested gene set with the trait, and not with other gene sets, meaning that the null hypothesis in this case is that the genes in the gene set are not associated with the trait [24, 25]. Therefore, for a competitive test, there should be data for the whole breadth of the genome, but this test cannot provide information regarding how strongly the gene set is associated with the trait [26]. We employ both methods for a comprehensive investigation into the neurobiological background of TS.

### MAGMA on raw genotypes

We ran MAGMA [23] on the individual-level genotype data using the aforementioned filtered gene set lists. MAGMA performs a three step analytic process. First it annotates the SNPs by assigning them to genes, based on their chromosomal location. Then it performs a gene prioritization step, which is used to perform the final gene set analysis step. We used a genomic window size of *±*10kb and the top 5 principal components as covariates to capture population structure. SNP to gene assignments were based on the NCBI 37.3 human gene reference build. The number of permutations required for the analysis was determined by MAGMA, using an adaptive permutation procedure leading to 11,263 permutations. MAGMA employs a family-wise error correction calculating a significance threshold of 0.00100496.

### Set-Based Association (SBA) test

We conducted SBA tests on the raw individual genotype data, as described in PLINK [24, 27] and adapted in a later publication [28]. This test relies on the assignment of individual SNPs to a gene, based on their position, and thus to a pathway, according to the NCBI 37.3 human gene reference build. After single marker association analysis, the top LD-independent SNPs from each set are retained and selected in order of decreasing statistical significance, and the mean of their association p-values is calculated. We permuted the case/control status, repeating the previous association and calculation steps described above, leading to the empirical p-value for each set. The absolute minimum number of permutations required for crossing the significance level is dictated by the number of gene sets tested. Testing for 45 gene sets requires at least 1000 permutations to produce significant findings. PLINK’s max(t) test recommends at least 64,000 permutations. We opted to increase the number of permutations to one million, the maximum that was computationally feasible, to maximize our confidence in the outcomes, given our large sample size.

We used logistic regression as the association model on the genotypes and the first five principal components as covariates on the genotype data to conduct the SBA test with the collected neural gene sets. Another repetition of this step was performed with a simple association test, to test for this method’s robustness to population structure. We proceeded to run the analysis on all samples, using all gene sets at a 10kb genomic window size, the first five principal components as covariates, and one million permutations. Since the permutations were performed on the phenotypic status of the samples, and only served as a method of association of the trait with the gene sets, we also corrected the results by defining the significance threshold through Bonferroni correction at 1.1*x* 10^*−*3^(0.05*/*45).

## Results

For the gene set association analysis, we ran PLINK’s self-contained set-based association method and MAGMA’s competitive association method, using the same 45 gene sets on the processed genotyped data of 3581 cases and 7682 ancestry-matched controls on a total of 236,248 SNPs. By performing both methods of analysis we aimed to obtain a global assessment of the gene sets’ relationship with TS.

MAGMA analysis identified one significant gene set (Figure 1), Cell Adhesion and Trans-synaptic Signaling (CATS), which achieved a nominal p-value of 6.2*x* 10^*−*5^ (permuted p-value of 0.0032). While the CATS gene set is comprised of 83 genes, MAGMA’s annotation step prioritized 72 of its genes for the gene set analysis. It involves 3290 variants that were reduced to 1627 independent variants in our data. Results were mainly driven by associations in the CDH26, CADM2 and OPCML genes as indicated by MAGMA gene-based analysis (Table 1). In the gene-based tests, CDH26 attained a p-value of 8.9526*x* 10^*−*6^, CADM2 a p-value of 4.6253*x* 10^*−*4^, and OPCML a p-value of 7.9851*x* 10^*−*4^, neither crossing the genome-wide significance threshold for gene tests (2.574*x* 10^*−*6^ calculated on 19,427 genes contained in the NCBI37.3 version of RefGene).

**Table 1:**
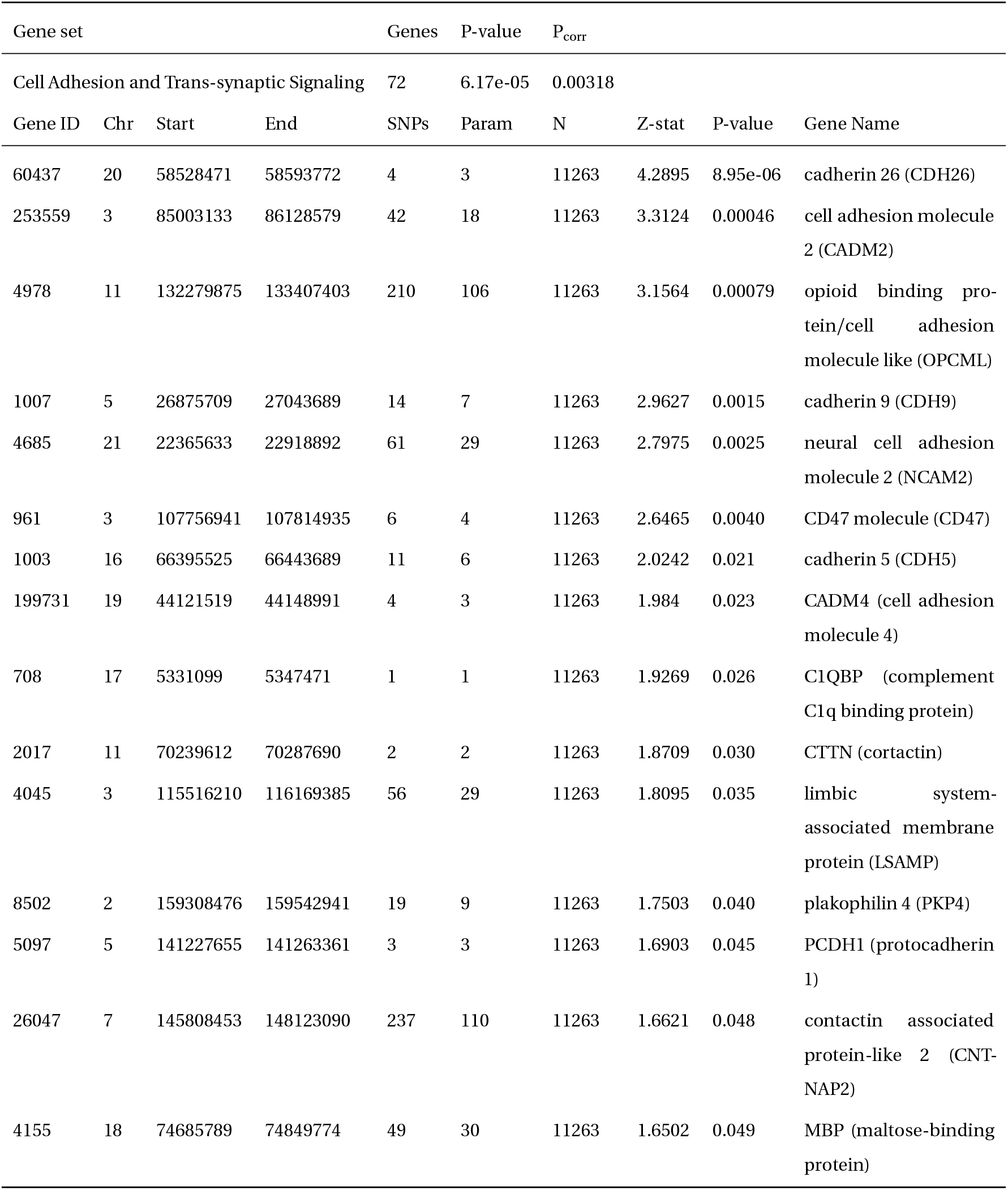
Statistically significant result of MAGMA gene set analysis. The Cell adhesion and transsynaptic signaling gene set achieved statistical significance. Genes within this set that achieved nominal significance with gene-based test implemented by MAGMA are also listed here. Gene ID refers to Entrez ID, Param to the number of SNPs used for the snp-wise analysis.

| Gene set |  |  |  | Genes | P-value | P<sub>corr</sub> |  |  |  |
| --- | --- | --- | --- | --- | --- | --- | --- | --- | --- |
| Cell Adhesion and Trans-synaptic Signaling |  |  |  | 72 | 6.17e-05 | 0.00318 |  |  |  |
| Gene ID | Chr | Start | End | SNPs | Param | N | Z-stat | P-value | Gene Name |
| 60437 | 20 | 58528471 | 58593772 | 4 | 3 | 11263 | 4.2895 | 8.95e-06 | cadherin 26 (CDH26) |
| 253559 | 3 | 85003133 | 86128579 | 42 | 18 | 11263 | 3.3124 | 0.00046 | cell adhesion molecule 2 (CADM2) |
| 4978 | 11 | 132279875 | 133407403 | 210 | 106 | 11263 | 3.1564 | 0.00079 | opioid binding protein/cell adhesion molecule like (OPCML) |
| 1007 | 5 | 26875709 | 27043689 | 14 | 7 | 11263 | 2.9627 | 0.0015 | cadherin 9 (CDH9) |
| 4685 | 21 | 22365633 | 22918892 | 61 | 29 | 11263 | 2.7975 | 0.0025 | neural cell adhesion molecule 2 (NCAM2) |
| 961 | 3 | 107756941 | 107814935 | 6 | 4 | 11263 | 2.6465 | 0.0040 | CD47 molecule (CD47) |
| 1003 | 16 | 66395525 | 66443689 | 11 | 6 | 11263 | 2.0242 | 0.021 | cadherin 5 (CDH5) |
| 199731 | 19 | 44121519 | 44148991 | 4 | 3 | 11263 | 1.984 | 0.023 | CADM4 (cell adhesion molecule 4) |
| 708 | 17 | 5331099 | 5347471 | 1 | 1 | 11263 | 1.9269 | 0.026 | C1QBP (complement C1q binding protein) |
| 2017 | 11 | 70239612 | 70287690 | 2 | 2 | 11263 | 1.8709 | 0.030 | CTTN (cortactin) |
| 4045 | 3 | 115516210 | 116169385 | 56 | 29 | 11263 | 1.8095 | 0.035 | limbic system-associated membrane protein (LSAMP) |
| 8502 | 2 | 159308476 | 159542941 | 19 | 9 | 11263 | 1.7503 | 0.040 | plakophilin 4 (PKP4) |
| 5097 | 5 | 141227655 | 141263361 | 3 | 3 | 11263 | 1.6903 | 0.045 | PCDH1 (protocadherin 1) |
| 26047 | 7 | 145808453 | 148123090 | 237 | 110 | 11263 | 1.6621 | 0.048 | contactin associated protein-like 2 (CNT-NAP2) |
| 4155 | 18 | 74685789 | 74849774 | 49 | 30 | 11263 | 1.6502 | 0.049 | MBP (maltose-binding protein) |

**Figure 1:**
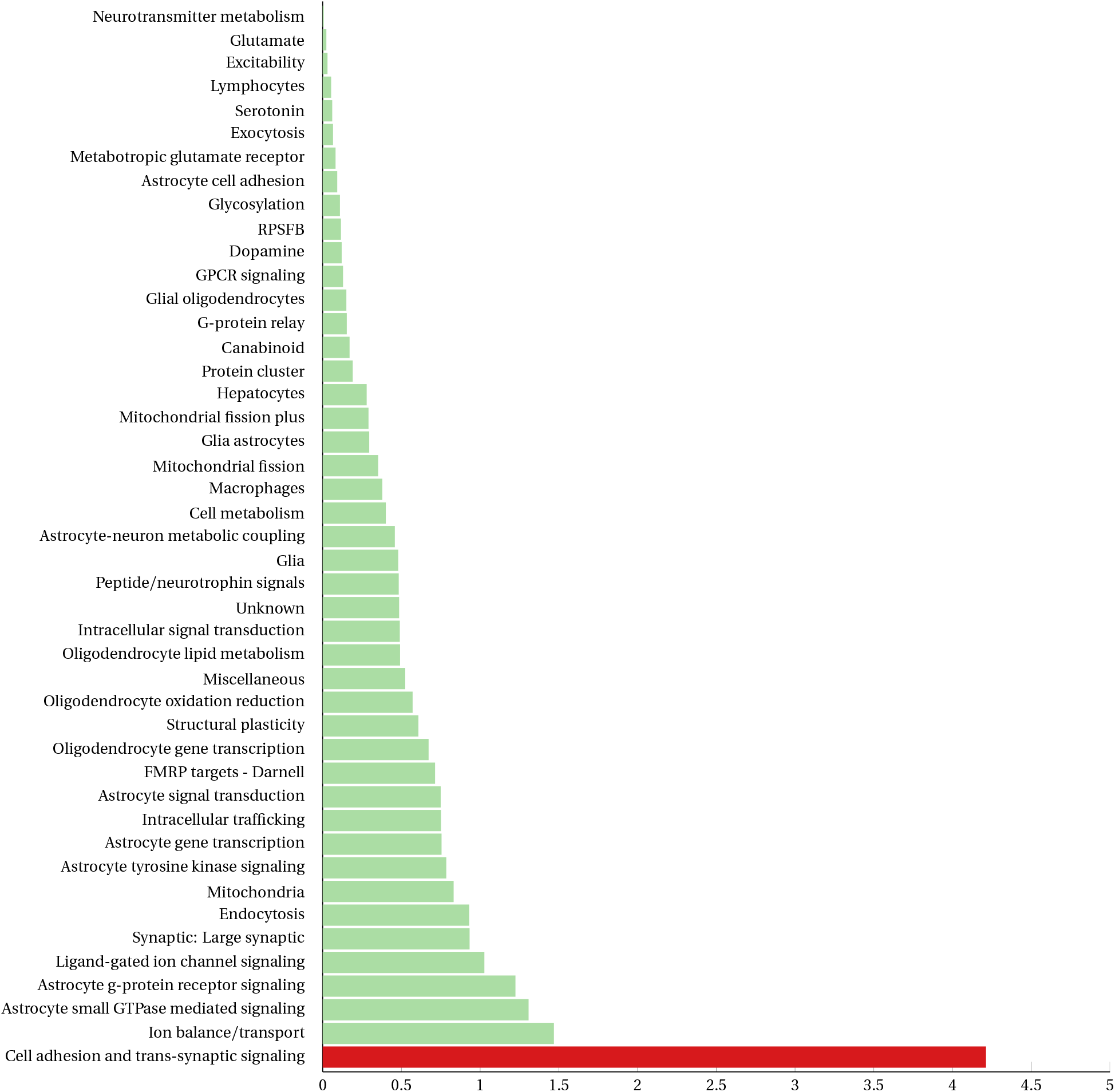
Results of gene set analysis as implemented by MAGMA. The gene set that crossed the significance threshold is depicted in red.

We next ran SBA, which conducts an initial single-marker association step before performing permutations to calculate empirical p-values for the gene sets. This association step is performed on the total number of variants that are associated with the genes involved in the gene sets, leading to a subset of 25,630 variants in our data, which are then filtered based on their LD. Analysis identified three gene sets as significant (Table 2), the Ligand-gated Ion Channel Signaling (LICS) (P: 2.67*x* 10^*−*4^), the Lymphocytic (P: 3.5*x* 10^*−*4^), and the Cell Adhesion and Trans-synaptic Signaling (CATS) (P: 1.07*x* 10^*−*3^). Detailed results for all the tested gene sets are shown in Figure 2.

**Table 2:** Statistically significant results of the SBA analysis. 3 pathways achieved significance. Association statistics for the top 5 SNPs driving the signal in each set are also shown. NSIG is the number of SNPs crossing the nominal significance threshold. EMP1 is the empirical p-value attained by the tested gene set. P is the p-value of the original single marker association, OR is the respective odds ratio. A1 is the minor allele and A2 the major allele. F_A and F_U are the frequencies of the minor allele in case and control samples respectively.

| Gene set |  |  |  | SNPs |  | NSIG | EMP1 |  |  |
| --- | --- | --- | --- | --- | --- | --- | --- | --- | --- |
| Ligand-gated Ion Channel Signaling |  |  |  | 683 |  | 66 | 0.000267 |  |  |
| Chr | SNP | BP | A1 | F_A | F_U | A2 | P | OR | Genes implicated |
| 4 | rs1391174 | 46072596 | T | 0.4892 | 0.4586 | C | 1.764e-05 | 1.131 | GABRG1(0) |
| 5 | rs9790873 | 45291514 | C | 0.1535 | 0.1335 | T | 5.621e-05 | 1.177 | HCN1(0) |
| 9 | rs2259639 | 101317401 | T | 0.2751 | 0.2982 | C | 0.0003612 | 0.8928 | GABBR2(0) |
| 9 | rs1930415 | 101238974 | T | 0.2218 | 0.2424 | C | 0.0007006 | 0.8908 | GABBR2(0) |
| 11 | rs949054 | 120795888 | C | 0.2241 | 0.2053 | T | 0.001281 | 1.118 | GRIK4(0) |
| Lymphocytes |  |  |  | 799 |  | 50 | 0.00035 |  |  |
| Chr | SNP | BP | A1 | F_A | F_U | A2 | P | OR | Genes implicated |
| 19 | rs16986092 | 55433696 | T | 0.1158 | 0.09473 | C | 1.093e-06 | 1.251 | NCR1(+9.257kb)\|NLRP7(-1.18kb) |
| 13 | rs1933437 | 28624294 | G | 0.4183 | 0.3871 | A | 8.482e-06 | 1.138 | FLT3(0) |
| 3 | rs2243123 | 159709651 | C | 0.2515 | 0.2759 | T | 0.0001167 | 0.8817 | IL12A(0)\|IL12A-AS1(0) |
| 7 | rs3801983 | 18683672 | C | 0.1928 | 0.2133 | T | 0.0003981 | 0.8808 | HDAC9(0) |
| 5 | rs2230525 | 66478626 | C | 0.08431 | 0.07127 | T | 0.0005641 | 1.2 | CD180(0) |
| Cell Adhesion and Trans-synaptic Signaling |  |  |  | 3290 |  | 292 | 0.00107 |  |  |
| Chr | SNP | BP | A1 | F_A | F_U | A2 | P | OR | Genes implicated |
| 20 | rs1002762 | 58580885 | G | 0.2305 | 0.2028 | A | 2.031e-06 | 1.178 | CDH26(0) |
| 21 | rs2826825 | 22762779 | G | 0.376 | 0.3487 | A | 6.698e-05 | 1.126 | NCAM2(0) |
| 11 | rs7925725 | 131449365 | C | 0.3709 | 0.3979 | A | 0.0001099 | 0.8921 | NTM(0) |
| 11 | rs12224080 | 131816849 | G | 0.09841 | 0.08353 | A | 0.0002519 | 1.198 | NTM(0) |
| 3 | rs6773575 | 77060574 | C | 0.0964 | 0.1126 | A | 0.000256 | 0.8407 | ROBO2(0) |

**Figure 2:**
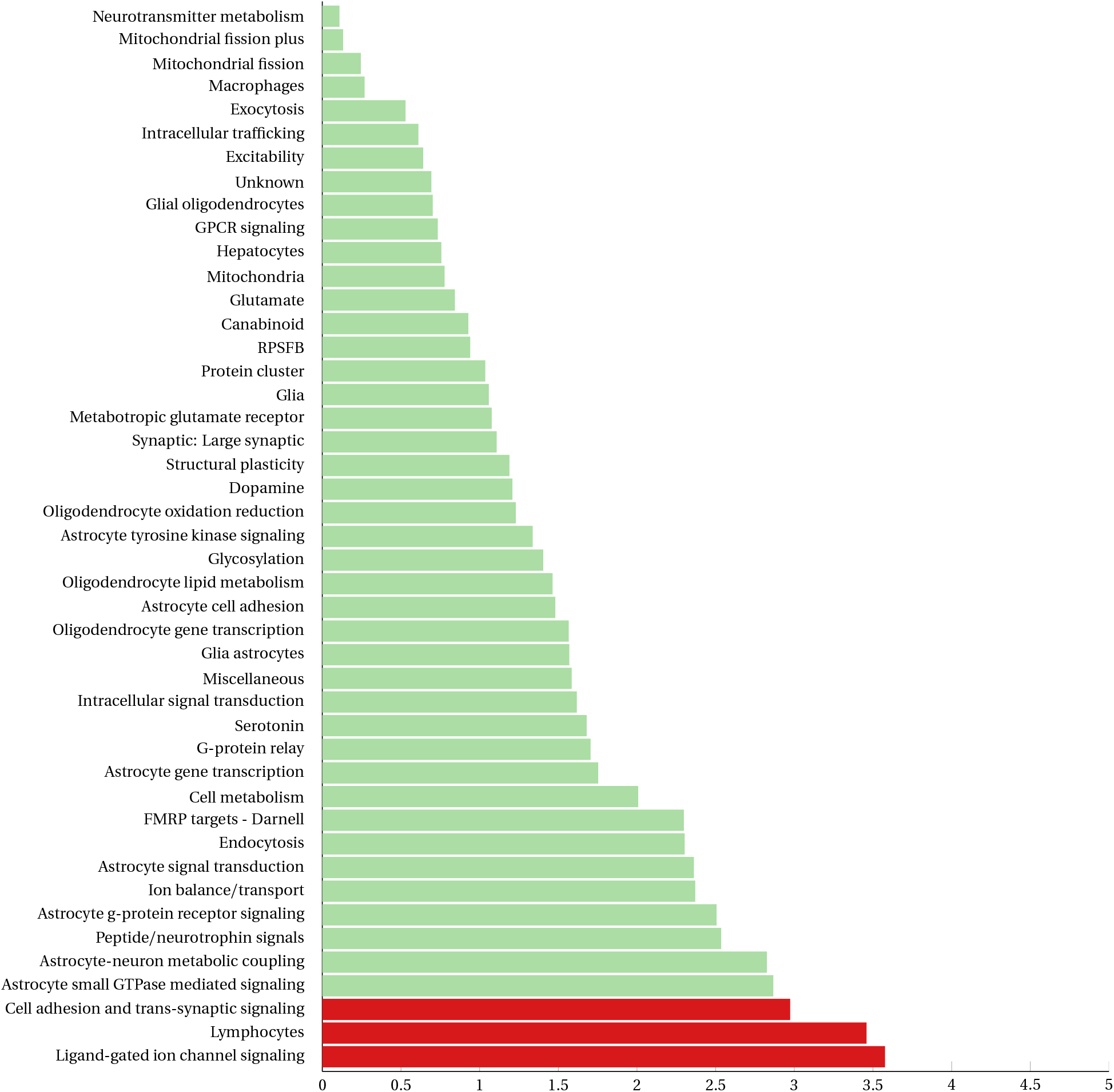
Results of gene set analysis as implemented by SBA. The gene sets that crossed the significance threshold are depicted in red.

The LICS gene set was the top scoring gene set, including 38 genes and involving 683 variants, 66 of which were associated with TS. The gene set’s signal was primarily driven by variants residing in the genes of the *γ*-aminobutyric acid receptors *GABRG1* and *GABBR2*, the *HCN1* channel gene and the glutamate receptor gene *GRIK4*. This signal was driven primarily by an association with SNP rs9790873, which is an eQTL for *HCN1* in tibial nerve, according to GTEx [29]. *GABBR2* is represented by two top SNPs, that are LD-independent, and removing either of those SNPs from the gene set did not cause the gene set to drop under the significance threshold.

The Lymphocytic gene set was the next top scoring gene set, including 143 genes that translated to 799 variants in our data, with 50 of these variants associated with TS. Its signal was driven by a missense variant inside the *FLT3* gene and an intergenic variant between *NCR1* and *NLRP7*, followed by *IL12A, HDAC9, CD180*. The rs1933437 SNP is the top variant for *FLT3*, and is a possibly damaging missense variant [30], located in the sixth exon of the *FLT3* gene leading to a p.Thr227Met mutation. It is a very common variant and the sixth exon appears to be less expressed than downstream exons. Given the tissues in which this eQTL affects *FLT3*’s expression, we tested the Lymphocytic gene set by removing *FLT3* from it, to identify whether the lymphocytic gene set association was biased by the presence of *FLT3*. After removing *FLT3*, the Lymphocytic gene set association statistic decreased slightly (P: 0.00012), driven mainly by *NCR1*/*NRLP7*.

The third significant gene set, CATS, consisted of 83 genes, including multiple large genes. CATS was identified by both SBA and MAGMA in our analyses, and both gene set approaches identified *CDH26* as the gene with the lowest p-value. Both SBA and MAGMA also identified *NCAM2, NTM* and *ROBO2* as strongly associated with TS, with *NTM* represented by two LD-independent SNPs. CATS’s top SNP, rs1002762, resides in the *CDH26* gene on chromosome 20, and is the top associated SNP in our data (P: 2.031*x* 10^*−*6^) with an odds-ratio of 1.178.

Notable results from the SBA also include the Astrocyte small GTPase mediated signaling (ASGMS) and the Astrocyte-neuron metabolic coupling (ANMC) gene sets, with a p-values slightly under the significance thresholds. These gene sets attained a p-value of 0.00137 and 0.001504 respectively.

## Discussion

Seeking to elucidate the neurobiology of TS, we present here the largest study to date aiming to interrogate the involvement of sets of genes that are related to neuronal and glial function in TS. We analyzed data from our recently performed TS GWAS and conducted two distinct types of testing, a competitive, regression-based test (MAGMA) and a self-contained, p-value combining test (SBA). Self-contained tests investigate for associations with a phenotype, while competitive tests compare a specific gene set against randomly generated gene sets. We employed both methods to perform a comprehensive investigation of the biological background of TS.

MAGMA’s regression-based algorithm has been reported to account for gene size biases, as can be also discerned by the variable sizes of the top genes. MAGMA’s top prioritized gene, *CDH26*, is represented by 4 SNPs in our data, *CADM2* by 42, while *OPCML* is represented by 210 SNPs, as it covers an extensive genomic region. We addressed such issues in SBA by setting a low *r* ^2^ threshold and conditioning on any LD-independent SNPs that resided on the same gene.

The gene sets used in our study come from a line of function-based analyses, aiming to investigate neurobiological mechanisms in neuropsychiatric disorders. A previous pathway analysis using individual level genotype data of the first wave TS GWAS identified genes involved in astrocyticneuron metabolic coupling, implicating astrocytes in TS pathogenesis [14]. In this study, we took advantage of the increased sample size of the second wave TS GWAS and the mechanics of the two distinct methods to identify gene sets associated with TS that provide a novel insight into the pathogenesis of TS, and substantiate the role of neural processes in this neuropsychiatric disorder.

The ANMC gene set that contains genes involved in carbohydrate metabolism in astrocytes was the single identified gene set in the previous pathway analysis study on TS [14], raising a hypothesis on a potential mechanism that involves altered metabolism of glycogen and glutamate/*γ*-aminobutyric acid in the astrocytes. In our study the ANMC gene set scored slightly under the significance threshold.

Here, analyzing a much larger sample size we identified 3 sets of genes as significantly associated to the TS phenotype. Among them the LICS gene set, which involves genes implicated in ion channel signaling through *γ*-aminobutyric acid and glutamate. Several genes in the LICS gene set have been previously implicated in neuropsychiatric phenotypes. *HCN1*, a hyperpolarization-activated cation channel involved in native pacemaker currents in neurons and the heart, has been significantly associated with schizophrenia and autism [31–33]. *GABRG1*, an integral membrane protein that inhibits neurotransmission by binding to the benzodiazepine receptor, has yielded mild associations with general cognitive ability [34] and epilepsy [35], while *GABBR2*, a g-protein coupled receptor that regulates neurotransmitter release, with schizophrenia [36] and post-traumatic stress disorder [37] in multiple studies. The GABA-ergic pathway has been previously implicated in TS, and recent advances showcased the possibility that a GABA-ergic transmission deficit can contribute towards TS symptoms[38]. *GRIK4*, encoding a glutamate-gated ionic channel, has shown associations with mathematical ability and educational attainment [39] and and weaker associations with attention-deficit hyperactivity disorder [40]. The *γ*-aminobutyric acid receptors and the HCN channel, are features of inhibitory interneurons [41] and also identified in the brain transcriptome of individuals with TS [42], adding to the evidence that the phenotype of TS could be influenced by an inhibitory circuit dysfunction, as has previously been proposed [43].

Individuals with TS are reported to present elevated markers of immune activation [42, 44]. Additionally, a number of studies have implicated neuroimmune responses with the pathogenesis of TS [45–47]. We investigated neuro-immune interactions by interrogating association to a gene set designed by Goudriaan et al [21] to study enrichment in lymphocytic genes. Indeed, our analysis yielded a statistically significant signal. The *FLT3* association coincides with the results of the second wave TS GWAS, in which *FLT3* was the only genome-wide significant hit [7]. *FLT3* and its ligand, *FLT3LG*, have a known role in cellular proliferation in leukemia, and have been found to be expressed in astrocytic tumors [48]. The rs1933437 variant in *FLT3* is an eQTL in the brain cortex and the cerebellum[29], and has also been implicated in the age at the onset of menarche [49]. Variants in *FLT3* have attained genome-wide significance in a series of studies focusing on blood attributes in populations of varying ancestry, and our current insights into its role are mostly based on these associations with blood cell counts, serum protein levels, hypothyroidism and autoimmune disorders [49–52].

*FLT3* could play a role in neuroinflammation as supported by its intriguing association with peripheral neuropathic pain. The inhibition of *FLT3* is reported to alleviate peripheral neuropathic pain (PNP) [53], a chronic neuro-immune condition that arises from aberrations in the dorsal root ganglia. Cytokines and their receptors have been at the epicenter of the neuro-immune interactions, with microglia contributing significantly to chronic phenotypes of such states [54]. *FLT3* is a critical component for neuro-immune interactions, especially in the case of the development and sustenance of the PNP phenotype. Interestingly, pain follows sex-specific routes, with glia having a prominent role for pain propagation in males, while females involve adaptive immune cells instead [55]. These, paired with previous evidence of glial involvement in TS [14], raise an interesting hypothesis for TS symptom sustenance, since *FLT3* has been shown to be critical for the chronicity of neuronal dysregulations [53].

FLT3 is expressed in the cerebellum and whole blood, while *FLT3*’s top variant, rs1933437, is an eQTL for *FLT3* on GTEx [29] in various brain tissues, such as the cortex, the cerebellum, the hypothalamus, the frontal cortex (BA9) and non-brain tissues, such as the skin, the pancreas and adipose tissues. In order to test the robustness of the lymphocytic association in our findings, we repeated the analysis after removing *FLT3* from the lymphocytic gene set. The p-value of the gene set decreased, but still remained significant, due to the association in the *NCR1/NLRP7* locus. Besides *FLT3*, the other genes included in this gene set are also quite intriguing to consider as potential candidates that could underlie the pathophysiology of TS. In the same vein with *FLT3*, common variants in NCR1 have also been significantly associated with blood protein levels [56]. HDAC9 has been significantly associated with androgenetic alopecia [49, 57], hair color [49], and ischemic stroke [58]. These seem to follow previous knowledge, given that genes involved in ischemic stroke have been identified as a common component between TS and ADHD [59], and that TS, similar to other neuropsychiatric disorders, demonstrates a distinct preference for males. CD180 has shown associations with general cognitive ability [34].

The CATS gene set involves many cadherins, with the top signals being in CDH26. CDH26 is a cadherin that regulates leukocyte migration, adhesion and activation, especially in the case of allergic inflammation [60]. Cell adhesion molecules have been consistently implicated in phenotypes pertaining to brain function, with the latest addition of the high confidence TS gene CELSR3, a flamingo cadherin, that was identified in a large scale de novo variant study for TS [12].

Most of the genes contained in the identified gene sets in this study are involved in cognitive performance, mathematical ability and educational attainment [39]. OPCML, CADM2, and ROBO2 have been implicated in neuromuscular and activity phenotypes, such as grip strength [61], physical activity [62], and body mass index [49]. ROBO2 has been associated with depression [63], expressive vocabulary in infancy [64], while CADM2 is associated to a multitude of phenotypes, including anxiety [63], risk taking behavior, smoking [65]. NTM displays similar patterns of pleiotropy, associated with smoking [49], myopia [57], hair color [66], anxiety [63], asperger’s syndrome [67], bipolar disorder with schizophrenia[68], and eating disorders [69]. NCAM2 and NTM, similarly to the lymphocytic genes, have been significantly associated with blood protein levels [70] and leukocyte count [49] respectively. Many of these phenotypes are known TS comorbidities, presenting themselves commonly or less commonly in TS cases, and other are related to functions that get impaired in TS symptomatology.

The CATS gene set was identified in both methods indicating the involvement of cell adhesion molecules in trans-synaptic signaling. Using genotypes with both methods as a means of identifying pathways instead of summary statistics, gave our study the edge of sample-specific linkage disequilibrium rather than relying on an abstract linkage disequilibrium pattern reference. Our current understanding for the regional structures of the genome and the cis effects of genomic organization will aid the refinement of these associations as well as help shape our understanding of the pleiotropic mechanisms in the identified loci potentially responsible for disease pathogenesis.

In conclusion, our analysis provides further support for the role of *FLT3* in TS, strengthens the possibility for the involvement of the GABA-ergic biological pathway in TS pathogenesis, and provides parallel insights into possible mechanisms underlying tic persistence, and a possible correlation with glial-derived neuro-immune phenotypes.

## Data Availability

Data available through the Psychiatric Genomics Consortium.

https://www.med.unc.edu/pgc/download-results/

## Conflict of interest

IM has participated in research funded by the Parkinson Foundation, Tourette Association, Dystonia Coalition, AbbVie, Biogen, Boston Scientific, Eli Lilly, Impax, Neuroderm, Prilenia, Revance, Teva but has no owner interest in any pharmaceutical company. She has received travel compensation or honoraria from the Tourette Association of America, Parkinson Foundation, International Association of Parkinsonism and Related Disorders, Medscape, and Cleveland Clinic, and royalties for writing a book with Robert rose publishers.

KMV has received financial or material research support from the EU (FP7-HEALTH-2011 No. 278367, FP7-PEOPLE-2012-ITN No. 316978), the German Research Foundation (DFG: GZ MU 1527/3-1), the German Ministry of Education and Research (BMBF: 01KG1421), the National Institute of Mental Health (NIMH), the Tourette Gesellschaft Deutschland e.V., the Else-Kroner-Fresenius-Stiftung, and GW, Almirall, Abide Therapeutics, and Therapix Biosiences and has received consultant’s honoraria from Abide Therapeutics, Tilray, Resalo Vertrieb GmbH, and Wayland Group, speaker’s fees from Tilray and Cogitando GmbH, and royalties from Medizinisch Wissenschaftliche Verlagsgesellschaft Berlin, Elsevier, and Kohlhammer; and is a consultant for Nuvelution TS Pharma Inc., Zynerba Pharmaceuticals, Resalo Vertrieb GmbH, CannaXan GmbH, Therapix Biosiences, Syqe, Nomovo Pharma, and Columbia Care.

BMN is a member of the scientific advisory board at Deep Genomics and consultant for Camp4 Therapeutics, Takeda Pharmaceutical and Biogen.

MMN has received fees for memberships in Scientific Advisory Boards from the Lundbeck Foundation and the Robert-Bosch-Stiftung, and for membership in the Medical-Scientific Editorial Office of the Deutsches Ärzteblatt. MMN was reimbursed travel expenses for a conference participation by Shire Deutschland GmbH. MMN receives salary payments from Life & Brain GmbH and holds shares in Life & Brain GmbH. All this concerned activities outside the submitted work.

MSO serves as a consultant for the Parkinson’s Foundation, and has received research grants from NIH, Parkinson’s Foundation, the Michael J. Fox Foundation, the Parkinson Alliance, Smallwood Foundation, the Bachmann-Strauss Foundation, the Tourette Syndrome Association, and the UF Foundation. MSO’s DBS research is supported by: NIH R01 NR014852 and R01NS096008. MSO is PI of the NIH R25NS108939 Training Grant. MSO has received royalties for publications with Demos, Manson, Amazon, Smashwords, Books4Patients, Perseus, Robert Rose, Oxford and Cambridge (movement disorders books). MSO is an associate editor for New England Journal of Medicine Journal Watch Neurology. MSO has participated in CME and educational activities on movement disorders sponsored by the Academy for Healthcare Learning, PeerView, Prime, QuantiaMD, WebMD/Medscape, Medicus, MedNet, Einstein, MedNet, Henry Stewart, American Academy of Neurology, Movement Disorders Society and by Vanderbilt University. The institution and not MSO receives grants from Medtronic, Abbvie, Boston Scientific, Abbott and Allergan and the PI has no financial interest in these grants. MSO has participated as a site PI and/or co-I for several NIH, foundation, and industry sponsored trials over the years but has not received honoraria. Research projects at the University of Florida receive device and drug donations.

DW receives royalties for books on Tourette Syndrome with Guilford Press, Oxford University Press, and Springer Press.

The rest of the authors declare that they have no conflict of interest.

## Acknowledgements

This research is co-financed by Greece and the European Union (European Social Fund-ESF) through the Operational Programme ≪Human Resources Development, Education and Lifelong Learning≫ in the context of the project “Reinforcement of Postdoctoral Researchers - 2nd Cycle” (MIS-5033021), implemented by the State Scholarships Foundation (IKY).

LKD was supported by grants from the National Institutes of Health including U54MD010722-04, R01NS102371, R01MH113362, U01HG009086, R01MH118223, DP2HD98859, R01DC16977, R01NS105-746, R56MH120736, R21HG010652, and RM1HG009034.

The members of the Tourette Association of America International Consortium for Genetics are Cathy L. Barr, James R. Batterson, Cheston Berlin, Cathy L. Budman, Danielle C. Cath, Giovanni Coppola, Nancy J. Cox, Sabrina Darrow, Lea K. Davis, Yves Dion, Nelson B. Freimer, Marco A. Grados, Erica Greenberg, Matthew E. Hirschtritt, Alden Y. Huang, Cornelia Illmann, Robert A. King, Roger Kurlan, James F. Leckman, Gholson J. Lyon, Irene A. Malaty, Carol A. Mathews, William M. McMahon, Benjamin M. Neale, Michael S. Okun, Lisa Osiecki, Danielle Posthuma, Mary M. Robertson, Guy A. Rouleau, Paul Sandor, Jeremiah M. Scharf, Harvey S. Singer, Jan Smit, Jae Hoon Sul, and Dongmei Yu.

The members of the Gilles de la Tourette Syndrome Genome-Wide Association Study Replication Initiative are Harald Aschauer, Csaba Barta, Cathy L. Budman, Danielle C. Cath, Christel Depienne, Andreas Hartmann, Johannes Hebebrand, Anastasios Konstantinidis, Carol A. Mathews, Kirsten R. Muller-Vahl, Peter Nagy, Markus M. Nöthen, Peristera Paschou, Renata Rizzo, Guy A. Rouleau, Paul Sandor, Jeremiah M. Scharf, Monika Schlögelhofer, Mara Stamenkovic, Manfred Stuhrmann, Fotis Tsetsos, Zsanett Tarnok, Tomasz Wolanczyk, and Yulia Worbe.

The members of the Tourette International Collaborative Genetics consortium are Lawrence W. Brown, Keun-Ah Cheon, Barbara J. Coffey, Andrea Dietrich, Thomas V. Fernandez, Blanca Garcia-Delgar, Donald L. Gilbert, Dorothy E. Grice, Julie Hagstrøm, Tammy Hedderly, Gary A. Heiman, Isobel Heyman, Pieter J. Hoekstra, Chaim Huyser, Young Key Kim, Young-Shin Kim, Robert A. King, Yun-Joo Koh, Sodahm Kook, Samuel Kuperman, Bennett L. Leventhal, Marcos Madruga-Garrido, Pablo Mir, Astrid Morer, Alexander Münchau, Kerstin J. Plessen, Veit Roessner, Eun-Young Shin, Dong-Ho Song, Jungeun Song, Jay A. Tischfield, A. Jeremy Willsey, and Samuel H. Zinner.

## Notes

### Funding Statement

FT was co-financed by Greece and the European Union (European Social Fund- ESF) through the Operational Programme «Human Resources Development, Education and Lifelong Learning» in the context of the project «Reinforcement of Postdoctoral Researchers - 2nd Cycle« (MIS-5033021), implemented by the State Scholarships Foundation (IKY).
LKD was supported by grants from the National Institutes of Health including U54MD010722-04, R01NS102371, R01MH113362, U01HG009086, R01MH118223, DP2HD98859, R01DC16977, R01NS105746, R56MH120736, R21HG010652, and RM1HG009034.

## References

1. Robertson MM, Cavanna AE, Eapen V. Gilles de la Tourette syndrome and disruptive behavior disorders: prevalence, associations, and explanation of the relationships. The Journal of neuropsychiatry and clinical neurosciences 27, 33–41 (2015).

2. Scharf JM, Miller LL, Gauvin CA, Alabiso J, Mathews CA, Ben-Shlomo Y. Population prevalence of Tourette syndrome: A systematic review and meta-analysis. Movement Disorders 30, 221–228 (2015).

3. Robertson MM, Eapen V, Cavanna AE. The international prevalence, epidemiology, and clinical phenomenology of Tourette syndrome: A cross-cultural perspective. Journal of Psychosomatic Research 67, 475–483 (2009).

4. Davis LK et al. Partitioning the Heritability of Tourette Syndrome and Obsessive Compulsive Disorder Reveals Differences in Genetic Architecture. PLoS Genetics. doi:10.1371/journal.pgen.1003864 (2013).

5. Robertson MM et al. Gilles de la Tourette syndrome. Nature Reviews Disease Primers 3, 16097 (2017).

6. Mataix-Cols D et al. Familial Risks of Tourette Syndrome and Chronic Tic Disorders: A Population-Based Cohort Study. JAMA psychiatry 72, 787–793 (2015).

7. Yu* D et al. Interrogating the genetic determinants of Tourette syndrome and other tic disorders through genome-wide association studies. American Journal of Psychiatry (2018).

8. Scharf JM et al. Genome-wide association study of Tourette’s syndrome. Molecular psychiatry 18, 721–8 (2013).

9. Abdulkadir M et al. Polygenic Risk Scores Derived From a Tourette Syndrome Genome-wide Association Study Predict Presence of Tics in the Avon Longitudinal Study of Parents and Children Cohort. Biological Psychiatry 85. Neurodevelopmental Alterations and Childhood Behavior, 298–304 (2019).

10. Huang A et al. Rare Copy Number Variants in NRXN1 and CNTN6 Increase Risk for Tourette Syndrome. Neuron 94. doi:10.1016/j.neuron.2017.06.010 (2017).

11. Willsey AJ et al. De Novo Coding Variants Are Strongly Associated with Tourette Disorder. Neuron 94, 486–499.e9 (2017).

12. Wang S et al. De Novo Sequence and Copy Number Variants Are Strongly Associated with Tourette Disorder and Implicate Cell Polarity in Pathogenesis. Cell Reports 24, 3441–3454.e12 (2018).

13. Ballard DH, Cho J, Zhao H. Comparisons of multi-marker association methods to detect association between a candidate region and disease. Genetic Epidemiology 34, 201–212 (2010).

14. De Leeuw C et al. Involvement of astrocyte metabolic coupling in Tourette syndrome pathogenesis. European Journal of Human Genetics 23, 1519–1522 (2015).

15. Turner S et al. Quality Control Procedures for Genome-Wide Association Studies. Current Protocols in Human Genetics, 1.19.1–1.19.18 (2011).

16. Lam M et al. RICOPILI: Rapid Imputation for COnsortias PIpeLIne. Bioinformatics. btz633. doi:10.1093/bioinformatics/btz633 (2019).

17. Price AL, Patterson NJ, Plenge RM, Weinblatt ME, Shadick NA, Reich D. Principal components analysis corrects for stratification in genome-wide association studies. Nature Genetics 38, 904–909 (2006).

18. Ruano D et al. Functional Gene Group Analysis Reveals a Role of Synaptic Heterotrimeric G Proteins in Cognitive Ability. American Journal of Human Genetics 86, 113–125 (2010).

19. Lips ES et al. Functional gene group analysis identifies synaptic gene groups as risk factor for schizophrenia. Molecular Psychiatry 17, 996–1006 (2012).

20. Duncan LE et al. Pathway analyses implicate glial cells in schizophrenia. PLoS ONE 9. doi:10.1371/journal.pone.0089441 (2014).

21. Goudriaan A et al. Specific glial functions contribute to Schizophrenia susceptibility. Schizophrenia Bulletin 40, 925–935 (2014).

22. Jansen A et al. Gene-set analysis shows association between FMRP targets and autism spectrum disorder. European Journal of Human Genetics 25, 863–868 (2017).

23. De Leeuw CA, Mooij JM, Heskes T, Posthuma D. MAGMA: Generalized Gene-Set Analysis of GWAS Data. PLoS Computational Biology 11. doi:10.1371/journal.pcbi.1004219 (2015).

24. Purcell S et al. PLINK: A Tool Set for Whole-Genome Association and Population-Based Linkage Analyses. The American Journal of Human Genetics 81, 559–575 (2007).

25. Goeman JJ, Buhlmann P. Analyzing gene expression data in terms of gene sets: methodological issues. Bioinformatics 23, 980–987 (2007).

26. Mooney MA, Wilmot B. Gene set analysis: A step-by-step guide. American Journal of Medical Genetics Part B: Neuropsychiatric Genetics 168, 517–527 (2015).

27. Chang CC, Chow CC, Tellier LC, Vattikuti S, Purcell SM, Lee JJ. Second-generation PLINK: rising to the challenge of larger and richer datasets. GigaScience 4, 7 (2015).

28. Skafidas E, Testa R, Zantomio D, Chana G, Everall IP, Pantelis C. Predicting the diagnosis of autism spectrum disorder using gene pathway analysis. Molecular Psychiatry 19, 504–510 (2014).

29. Carithers LJ et al. A Novel Approach to High-Quality Postmortem Tissue Procurement: The GTEx Project. Biopreservation and Biobanking 13, 311–319 (2015).

30. Karczewski KJ et al. Variation across 141,456 human exomes and genomes reveals the spectrum of loss-of-function intolerance across human protein-coding genes. bioRxiv. doi:10.1101/531210 (2019).

31. Pardiñas AF et al. Common schizophrenia alleles are enriched in mutation-intolerant genes and in regions under strong background selection. Nature genetics 50, 381–389 (2018).

32. Autism Spectrum Disorders Working Group of The Psychiatric Genomics Consortium. Meta-analysis of GWAS of over 16,000 individuals with autism spectrum disorder highlights a novel locus at 10q24.32 and a significant overlap with schizophrenia. Molecular Autism 8, 21 (2017).

33. Okbay A et al. Genome-wide association study identifies 74 loci associated with educational attainment. Nature 533, 539–542 (2016).

34. Davies G et al. Study of 300,486 individuals identifies 148 independent genetic loci influencing general cognitive function. Nature Communications 9, 2098 (2018).

35. International League Against Epilepsy Consortium on Complex Epilepsies. Genetic determinants of common epilepsies: a meta-analysis of genome-wide association studies. The Lancet Neurology 13, 893–903 (2014).

36. Ikeda M et al. Genome-Wide Association Study Detected Novel Susceptibility Genes for Schizophrenia and Shared Trans-Populations/Diseases Genetic Effect. Schizophrenia bulletin. doi:10.1093/schbul/sby140 (2018).

37. Xie P, Kranzler HR, Yang C, Zhao H, Farrer LA, Gelernter J. Genome-wide Association Study Identifies New Susceptibility Loci for Posttraumatic Stress Disorder. Biological Psychiatry 74, 656–663 (2013).

38. Puts NAJ et al. Reduced GABAergic inhibition and abnormal sensory symptoms in children with Tourette syndrome. Journal of Neurophysiology 114, 808–817 (2015).

39. Lee JJ et al. Gene discovery and polygenic prediction from a genome-wide association study of educational attainment in 1.1 million individuals. Nature genetics 50, 1112–1121 (2018).

40. Yang L et al. Polygenic transmission and complex neuro developmental network for attention deficit hyperactivity disorder: genome-wide association study of both common and rare variants. American journal of medical genetics. Part B, Neuropsychiatric genetics : the official publication of the International Society of Psychiatric Genetics 162B, 419–430 (2013).

41. Kelsom C, Lu W. Development and specification of GABAergic cortical interneurons. Cell & bioscience 3, 19 (2013).

42. Lennington JB et al. Transcriptome Analysis of the Human Striatum in Tourette Syndrome.Biological Psychiatry 79, 372–382 (2016).

43. Rapanelli M, Frick LR, Pittenger C. The Role of Interneurons in Autism and Tourette Syndrome.Trends in Neurosciences 40, 397–407 (2017).

44. Krause DL, Müller N. The Relationship between Tourette’s Syndrome and Infections. The open neurology journal 6, 124–8 (2012).

45. Ercan-Sencicek AG et al. L-Histidine Decarboxylase and Tourette’s Syndrome. New England Journal of Medicine 362, 1901–1908 (2010).

46. Castellan Baldan L et al. Histidine Decarboxylase Deficiency Causes Tourette Syndrome: Parallel Findings in Humans and Mice. Neuron 81, 77–90 (2014).

47. Alexander J et al. Targeted Re-Sequencing Approach of Candidate Genes Implicates Rare Potentially Functional Variants in Tourette Syndrome Etiology. Frontiers in Neuroscience 10, 428 (2016).

48. Eßbach C et al. Abundance of Flt3 and its ligand in astrocytic tumors. OncoTargets and therapy 6, 555–61 (2013).

49. Kichaev G et al. Leveraging Polygenic Functional Enrichment to Improve GWAS Power. American journal of human genetics 104, 65–75 (2019).

50. Astle WJ et al. The Allelic Landscape of Human Blood Cell Trait Variation and Links to Common Complex Disease. Cell 167, 1415–1429.e19 (2016).

51. Jain D et al. Genome-wide association of white blood cell counts in Hispanic/Latino Americans: the Hispanic Community Health Study/Study of Latinos. Human molecular genetics 26, 1193–1204 (2017).

52. Kanai M et al. Genetic analysis of quantitative traits in the Japanese population links cell types to complex human diseases. Nature genetics 50, 390–400 (2018).

53. Rivat C et al. Inhibition of neuronal FLT3 receptor tyrosine kinase alleviates peripheral neuropathic pain in mice. Nature Communications 9, 1042 (2018).

54. Marchand F, Perretti M, McMahon SB. Role of the Immune system in chronic pain. Nature Reviews Neuroscience 6, 521–532 (2005).

55. Sorge RE et al. Different immune cells mediate mechanical pain hypersensitivity in male and female mice. Nature Neuroscience 18, 1081–1083 (2015).

56. Sun BB et al. Genomic atlas of the human plasma proteome. Nature 558, 73–79 (2018).

57. Pickrell JK, Berisa T, Liu JZ, Ségurel L, Tung JY, Hinds DA. Detection and interpretation of shared genetic influences on 42 human traits. Nature genetics 48, 709–17 (2016).

58. Malik R et al. Multiancestry genome-wide association study of 520,000 subjects identifies 32 loci associated with stroke and stroke subtypes. Nature genetics 50, 524–537 (2018).

59. Tsetsos F et al. Meta-analysis of tourette syndrome and attention deficit hyperactivity disorder provides support for a shared genetic basis. Frontiers in Neuroscience 10. doi:10.3389/fnins.2016.00340 (2016).

60. Caldwell JM et al. Cadherin 26 is an alpha integrin-binding epithelial receptor regulated during allergic inflammation. Mucosal Immunology 10, 1190–1201 (2017).

61. Tikkanen E et al. Biological Insights Into Muscular Strength: Genetic Findings in the UK Biobank. Scientific reports 8, 6451 (2018).

62. Klimentidis YC et al. Genome-wide association study of habitual physical activity in over 377,000 UK Biobank participants identifies multiple variants including CADM2 and APOE. International Journal of Obesity 42, 1161–1176 (2018).

63. Nagel M, Watanabe K, Stringer S, Posthuma D, van der Sluis S. Item-level analyses reveal genetic heterogeneity in neuroticism. Nature Communications 9, 905 (2018).

64. St Pourcain B et al. Common variation near ROBO2 is associated with expressive vocabulary in infancy. Nature communications 5, 4831 (2014).

65. Clifton EAD et al. Genome-wide association study for risk taking propensity indicates shared pathways with body mass index. Communications biology 1, 36 (2018).

66. Morgan MD et al. Genome-wide study of hair colour in UK Biobank explains most of the SNP heritability. Nature communications 9, 5271 (2018).

67. Salyakina D et al. Variants in several genomic regions associated with asperger disorder. Autism research : official journal of the International Society for Autism Research 3, 303–10 (2010).

68. Wang KS, Liu XF, Aragam N. A genome-wide meta-analysis identifies novel loci associated with schizophrenia and bipolar disorder. Schizophrenia research 124, 192–9 (2010).

69. Cornelis MC et al. A genome-wide investigation of food addiction. Obesity (Silver Spring, Md.) 24, 1336–41 (2016).

70. Emilsson V et al. Co-regulatory networks of human serum proteins link genetics to disease. Science (New York, N.Y.) 361, 769–773 (2018).

